# A multiscale mathematical model integrating viral load improves reconstruction of transmission chains and epidemiological parameters in household studies

**DOI:** 10.64898/2026.10.18.26364899

**Authors:** Nathanaël Hozé, Jérémie Guedj, Lulla Opatowski

**Affiliations:** Université Paris Cité and Université Sorbonne Paris Nord, Inserm, IAME, F-75018 Paris, France; Institut Pasteur, Université Paris Cité, Epidemiology and Modeling of Antibiotic Evasion (EMAE) Unit, Paris, France; Université Paris-Saclay, Université de Versailles Saint-Quentin-en-Yvelines, Inserm U1018, CESP, Anti-Infective Evasion and Pharmacoepidemiology Team, Montigny-Le-Bretonneux, France

## Abstract

Household studies combined with mathematical modelling are central to inference of respiratory pathogen transmission, but they usually rely on symptom onset or follow-up, final infection status, or qualitative PCR results. Quantitative viral-load measurements, although directly derived from PCR assays, are rarely used to reconstruct individual infection trajectories and transmission links.

We developed a Bayesian modeling framework integrating within-host viral dynamics and household transmission processes. Using simulated household outbreaks, we compared three observation designs: based on time of symptom onset, longitudinal qualitative PCR testing, and longitudinal quantitative viral load sampling. We evaluated their ability to recover unbiased infection times, transmission chains, generation intervals, incubation periods, transmission probabilities, and individual infectiousness profiles.

Quantitative viral-load improved transmission chains reconstruction compared with symptom onset and qualitative PCR designs. Across transmission-probability settings, perfect reconstruction of transmission chains reached 87-93%, compared with 46-57% using symptom-onset data and 68-78% using qualitative PCR. The correct number of generations of transmission was recovered in 94% of households. Variability in generation intervals, incubation periods, and transmission probabilities were recovered, whereas simpler designs mainly recovered average quantities and failed to capture inter-individual heterogeneity.

These findings support integrating longitudinal viral-load measurements into household transmission studies to improve inference of transmission dynamics and individual infectiousness.

**Author Summary:** Respiratory viruses often spread within households. Most household studies rely on symptom onset or simple positive or negative PCR results, which provide limited information about how infectious each individual was over time. It is therefore difficult to determine exactly when each person became infected and who infected whom. We developed a computational framework that combines individual viral-load trajectories with a household transmission model to reconstruct infection times, transmission chains, and key epidemiological quantities. Using simulated household outbreaks, we compared three types of study data: symptom onset only, repeated qualitative PCR testing, and repeated quantitative viral-load measurements. We found that quantitative viral-load data substantially improved reconstruction of transmission chains and recovered variation in infectiousness between individuals, whereas symptom-based and qualitative PCR designs mainly recovered average quantities and missed important heterogeneity. These results suggest that collecting longitudinal quantitative viral-load measurements can make household transmission studies more informative. More broadly, our framework shows how within-host biological measurements can be integrated with between-host transmission models to improve inference of infectious disease dynamics.

## Introduction

Household studies have been instrumental in estimating key epidemiological parameters, including secondary attack rates, incubation periods, intervention effects, and heterogeneity in individual infectiousness, for respiratory infections such as influenza [1,2], respiratory syncytial virus [3], and SARS-CoV-2 [4-6]. Their strength lies in the relatively closed household environment, where contacts are frequent and follow-up can be standardized. However, infection times and transmission links are rarely observed directly, making it difficult to reconstruct who infected whom and when transmission occurred.

Most household analyses infer infection timing from symptom onset or final infection status. This approach is imprecise for pathogens with variable incubation periods and for infections that remain asymptomatic. Prospective studies with repeated PCR sampling can refine infection timing, particularly among asymptomatic individuals [7,8], but qualitative PCR results provide limited information on differences in infectiousness between infected individuals.

Viral load, the quantity of virus measured in respiratory samples, has been associated with infectiousness and transmission risk [9,10]. Longitudinal quantitative PCR measurements can therefore provide information on individual viral trajectories and on the timing of infectiousness. Yet such measurements are rarely integrated into household transmission models, and to our knowledge, no household transmission modelling framework has jointly integrated longitudinal individual viral-load trajectories, within-host viral kinetics, and between-host transmission inference to reconstruct transmission chains and epidemiological parameter distributions.

In this study, we investigated to what extent accounting for individual viral-load trajectories improves reconstruction of household transmission. We developed a Bayesian multiscale modelling framework that combines within-host viral kinetics [11,12], probabilistic infectiousness profiles, and between-host household transmission. The present implementation was designed to mimic SARS-CoV-2 infection. The framework can be adapted to other respiratory pathogens by modifying the viral-kinetic, observation, and transmission models. We evaluated the framework exclusively using simulated household outbreaks. We used this framework to compare three observation designs (Figure 1): (i) symptom-onset-based data only, (ii) repeated qualitative PCR testing, and (iii) longitudinal quantitative PCR sampling and to assess their ability to reconstruct infection times, transmission chains, generation intervals, incubation periods, transmission probabilities, and heterogeneity in infectiousness.

**Figure 1.**
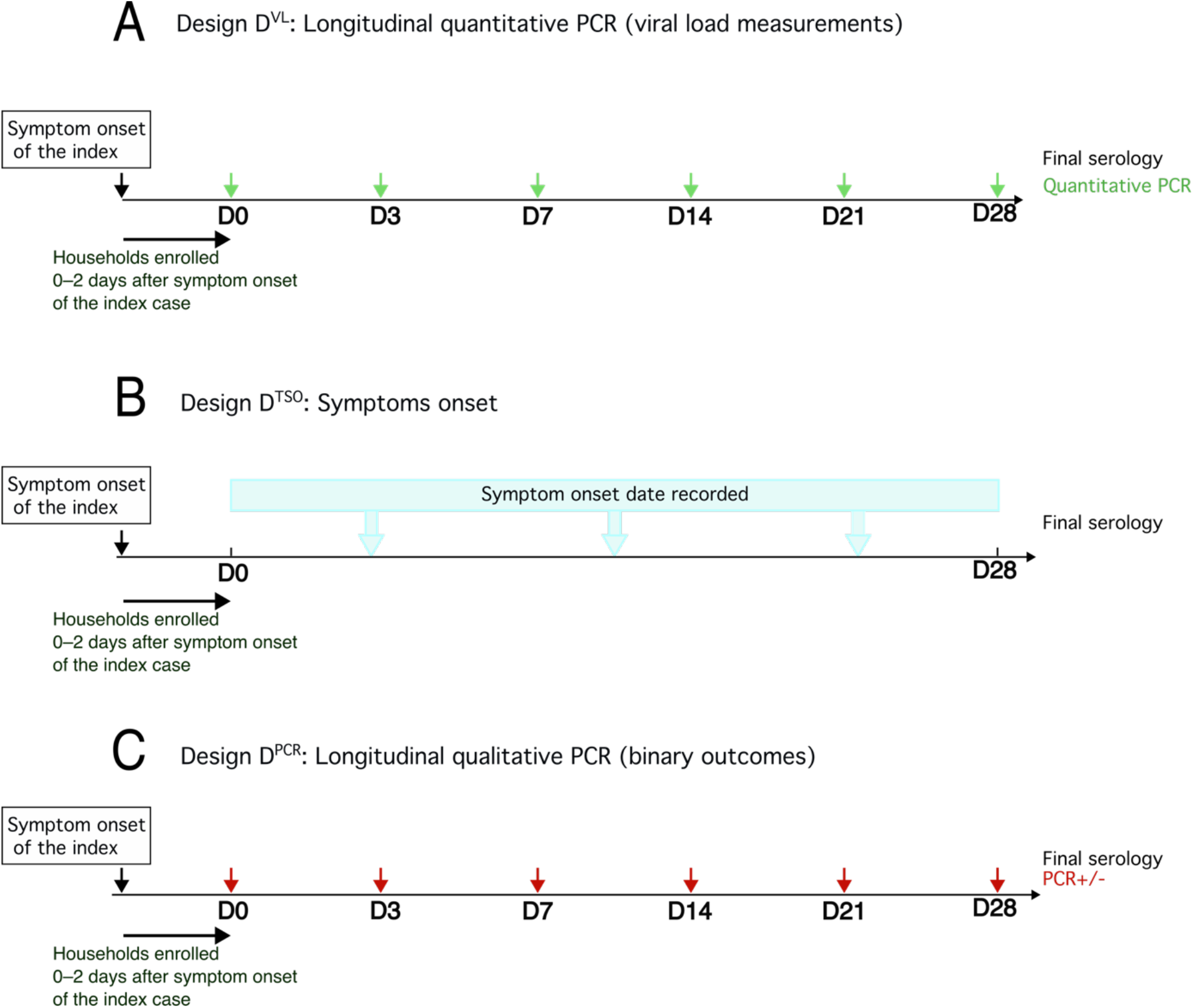
Study designs for household transmission inference. Schematic representation of the three household study designs compared in this study. In all designs, households are enrolled between 0 and 2 days after symptom onset in the index case and followed for 28 days. D0 corresponds to the time of enrollment. Infection status is determined by serology at the end of the study. **(A)** *D*^*VL*^: longitudinal quantitative viral-load design, with quantitative PCR sampled at D0, D3, D7, D14, D21, and D28 for all household members and symptom onset recorded for the index case only. (**B**) *D*^*TSO*^: Symptom-based design. Symptom onset dates are recorded for all individuals. No virological measurements are collected. (**C**) *D*^*PCR*^: Longitudinal qualitative PCR design with the same sampling times as *D*^*VL*^ but binary positive/negative PCR results relative to the detection threshold.

## Methods

### Overview of the simulation-inference framework

We first simulated household outbreaks using individual viral-load trajectories and a viral-load-dependent transmission model. We then generated observations under three designs with decreasing virological information and fitted the corresponding Bayesian inference models. Performance was assessed by comparing posterior estimates of infection times, transmission chains, epidemiological parameter distributions, and infectiousness profiles with the simulated truth.

### Viral dynamics model

A phenomenological viral load model was developed, accounting for individual variability, and set to mimic SARS-CoV-2 infection dynamics at the individual level (Figure 2A). In the model, each infected individual trajectory of viral load V(t) followed a piecewise linear model in log-scale [11], with initialization at -2 log_10_ copies/mL at infection and linear increase until reaching the limit of detection (LOD, 2 log_10_ copies/mL). Viral load *V(t)* then increased for a proliferation phase of duration *T*_*P*_ to a peak value *V*_*P*_ (relative to the LOD). Following the peak, the viral load decreases during a clearance phase *T*_*U*_ until it reaches the LOD again. By construction, the limit of detection is attained after a time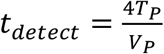. The time to detection is 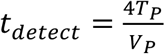, representing the interval between infection and when the LOD is first reached (Fig. 2A).

**Figure 2.**
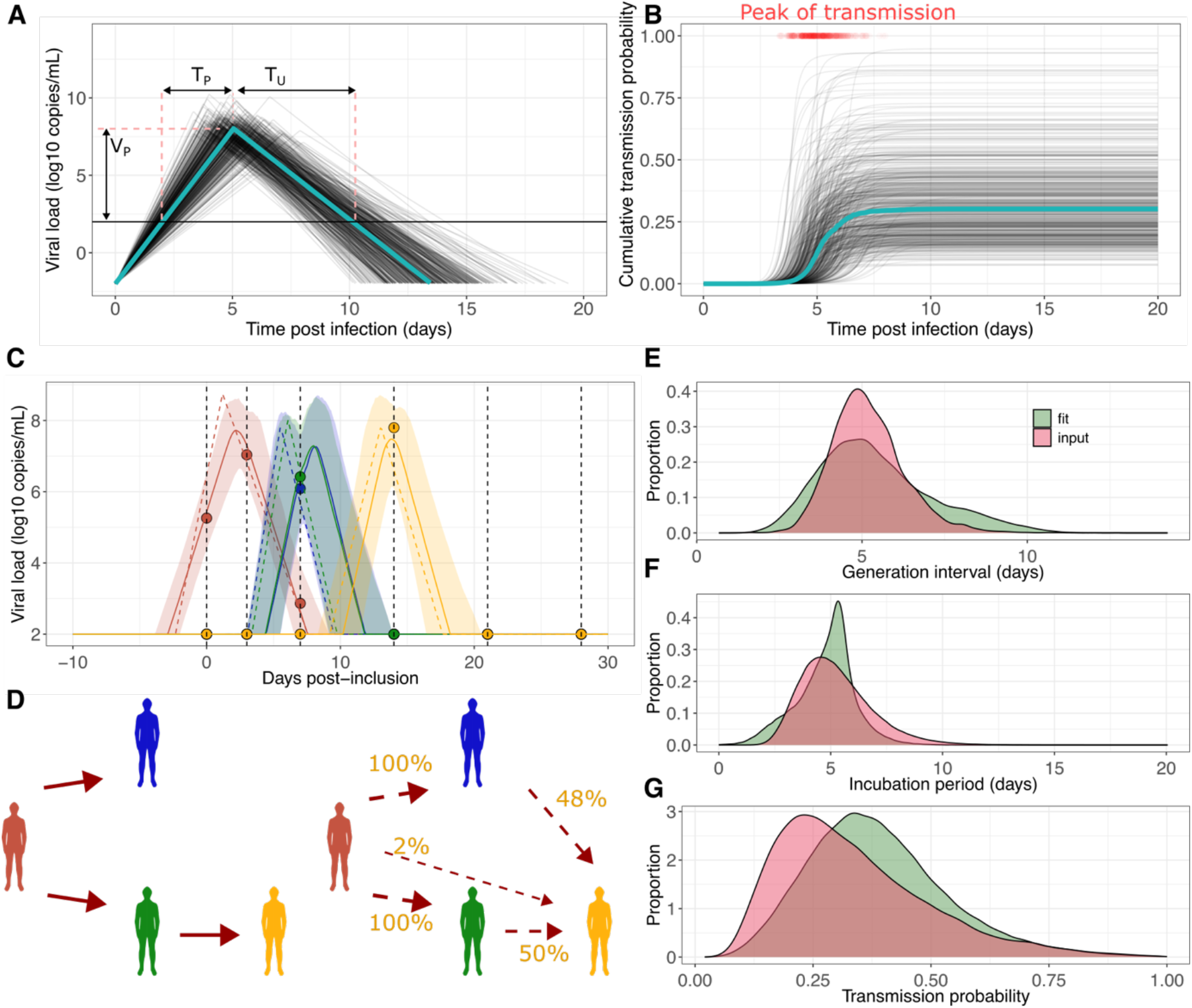
Linking individual viral dynamics to transmission. (**A**) Individual viral load trajectories for 500 simulated individuals, showing log viral load over time post infection based on individual parameters. The horizontal black line represents the detection limit (2 log10 copies/mL). The blue curve depicts viral dynamics using population-average parameters. (**B**) Individual cumulative transmission probability, with the blue curve representing population-average values and red points indicating the time of maximal transmission potential for each individual. (**C**) Reconstructed viral load trajectories for a representative household with four infected individuals under *D*^*VL*^. The true viral load trajectories are shown as dashed lines, observed viral load measurements as dots, and the posterior mean trajectory as solid lines with 95% credible intervals (shaded envelopes). Vertical dashed lines indicate sampling times. Reconstructed transmission chain across 1,500 posterior draws. In this household, the index case (red) infected blue and green contacts, and green subsequently infected yellow (true chain, left). Chain reconstruction after inference on the collected data (reconstructed chain, dashed, right) shows that the first transmissions are almost perfectly retrieved. The infector of the yellow individual is correctly inferred to be the green one in half of the simulated chains, the other half are blue and red only marginally. (**E**) Reconstruction of the chains of transmission and the time of infection allows estimating the generation interval. The posterior distribution of the estimated generation interval (red) is compared with the true distribution of the generation interval used to simulate the data (green). (**F**) Compared distributions of the input and estimated incubation period. (**G**) Compared distributions of the input and estimated infection probability. The reference simulation used *P*_*tr*_ = 0.30, *α* = 1, a mean generation interval of five days, a mean incubation period of 5 days.

Symptom onset was simulated as a log-normally distributed delay following infection. Individual parameters were drawn from lognormal distributions (Table S1).

### Household transmission

The within-host dynamic model was plugged into a between-host transmission model within households.

#### Model of transmission between two individuals

A dose-response model was considered to describe the relationship between the viral load and the transmission probability from an infected individual. The instantaneous hazard of transmission from an infectious individual to a susceptible individual is governed by the transmission-strength parameter *m*, and is given by

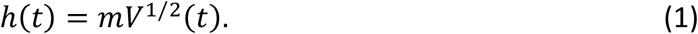

We assumed continuous and homogeneous contact between individuals, enabling the probability of transmission to be calculated by integrating over the entire period of contact. The density of transmission at time *t* is

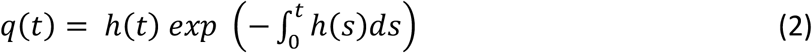

The cumulate transmission probability is

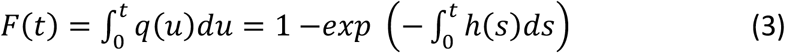

and the overall probability of transmission to a susceptible individual is given by:

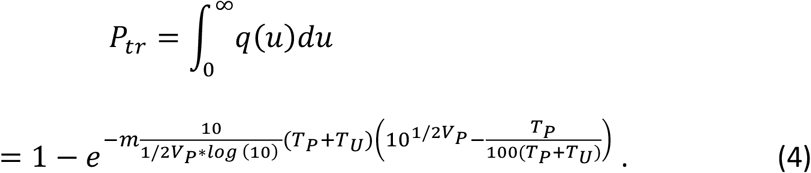

Conditional on transmission occurring, the transmission time density is *q*(*t*)/*P*_*tr*_. The overall transmission probability *P*_*tr*_ is the probability that this individual infects one continuously exposed individual over the infectious period. We calibrated *m* in a two-person household using population-level viral dynamics for the index case, so *P*_*tr*_ represents the cumulative infection probability over the infectious period. It therefore reflects cumulative exposure, not per-contact transmissibility or R0, which requires additional assumptions about contact frequency and infectiousness duration to be made. The overall transmission probability *P*_*tr*_ differs from the secondary attack rate (SAR), defined here as the average proportion of contacts infected across households, because the SAR also depends on household size.

#### Transmission within household

When several infectious individuals are present in a household, the household hazard of transmission is defined as the sum of individual hazards of transmission. Household size was accounted for using a scaling parameter *α*, which controls how the effective exposure to each infectious household member changes with the number of household contacts: *α* = 1 corresponds to frequency-dependent transmission, whereas *α* = 0 corresponds to density-dependent transmission. Specifically, in a household of size *n*, the force of infection *λ*_*s*_(*t*) experienced by susceptible individual *s* is

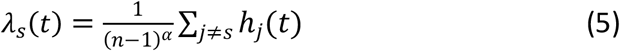

where *h*_*j*_(*t*) are the individual hazards of transmission which depends on the characteristics of infectious individuals *j* defined by equation (1).

#### The generation interval

The generation interval represents the time between infection of a primary case and infection of a secondary case. Using the probability density of infection over time, the mean generation interval is defined as

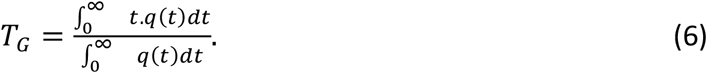

However, when individual-level parameters vary, this expression cannot be reported directly, as these parameters influence the value of *T*_*G*_. In such cases, the overall generation interval must be weighted by the probability that transmission occurs. Notably, individuals with higher transmissibility (e.g., larger viral peak values) tend to transmit infection earlier. We therefore reported the distribution of generation intervals weighted by the transmission probability (Figure 2E).

### Parameters used for simulations

We used parameter values chosen to mimic SARS-CoV-2-like viral kinetics and household transmission (Table S1). Individual viral-dynamics parameters were drawn from lognormal distributions. The transmission-strength parameter *m* was calibrated so that an individual with population-average viral dynamics had a cumulative probability of transmission to one susceptible household member equal to *P*_*tr*_. The main analysis used *P*_*tr*_ = 0.30, and additional simulations considered *P*_*tr*_ = 0.15 and *P*_*tr*_ = 0.45 to represent lower and higher transmission-probability settings consistent with the range for respiratory infections in households [13,14]. In the simulations, *m* was calibrated for the trajectory defined by the population-level viral-kinetic parameters to *m* = 1.4 10^−5^, *m* = 3.1 10^−5^ and *m* = 5.2 10^−5^ for *P*_*tr*_ = 0.15, 0.30, 0.45, respectively, and was fixed across individuals. The corresponding mean transmission probabilities across heterogeneous individuals were 0.17, 0.33, 0.47, respectively.

Therefore, heterogeneity in infectiousness arose from individual and temporal differences in viral load. This generated heterogeneous infectiousness profiles and transmission events concentrated around the period of high viral load (Figure 2B).

Household sizes were sampled from the French demographic distribution, mainly including households of two to six individuals [15] (median = 3, interquartile range: 2-4). The main simulations used frequency-dependent household transmission with alpha = 1, so that the probability of transmission to a given household member decreased as household size increased. Each simulation replicate included 100 households followed for 28 days.

### Observations and study designs

To evaluate the impact of data availability and more generally how study design strategies influence inference, we considered three observation designs, denoted *D*^*TSO*^, *D*^*PCR*^ and *D*^*VL*^ reflecting increasing levels of information. In all designs, households were enrolled 0-2 days after symptom onset in the index case and followed for 28 days, with final infection status determined at the end of follow-up.

*D*^*VL*^ **: Longitudinal quantitative viral-load design**. In the most informative design, individuals underwent PCR testing at days 0, 3, 7, 14, 21 and 28, and PCR measurements are quantitative, providing viral load measurements over time (Fig. 1A). Symptom onset was recorded for the index case only.

*D*^*TSO*^**: Symptoms**, uses only times of symptom onset for all participants, without virological measurements (Fig. 1B).

*D*^*PCR*^ **: Longitudinal qualitative PCR testing**. In this design, individuals are sampled at the same time points as in Design *D*^*VL*^, but observations were retained only as binary PCR detection status, positive or negative relative to the detection threshold (Figure 1C). Symptom onset was recorded for the index case only.

### Parameter estimation

In the following, we noted *N*_G_ the number of households. We noted *y*_*i,j,k*_ the observed viral loads, where the subscript *i* identifies the individual, *j* is the indexed time, in a household indexed by *k*. In a given household, *N*_K_ is the number of individuals, *N*_*I,k*_ being infected and *N*_*k*_− *N*_*I,k*_ non infected. We wrote *θ* the parameter vector, which is comprised of the transmission parameter *m*, the observation noise *σ*, and the individual parameter vector *θ* _*i,k*_ = (*T*_*I,i,k*_ ,*T*_*P,i,k*_*T*_*U,i,k*_, *V*_*P,i,k*_) that characterize the individual viral load dynamics. We wrote *V*_*i,i,k*_ the real viral load generated by the individual parameters.

#### Hierarchical structure of the model

We developed a hierarchical Bayesian framework to jointly estimate the within- and between-host parameters. The likelihood combines three components: the observation likelihood, the population distribution of individual viral-kinetic parameters, and the household infection likelihood. The approach is data-dependent and needs to be adapted with the design. We first give the likelihood when viral load is measured as in the quantitative and qualitative viral data in *D*^*VL*^ and *D*^*PCR*^. The individual parameters are treated as a latent variable and in the log-likelihood

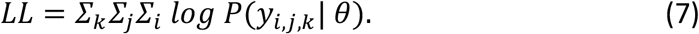

The elements of the likelihood can be written as

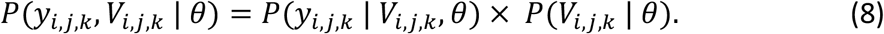

The first term on the right-hand side characterizes the probability of the observations conditional on the latent viral-load trajectories. The second term is the probability of the latent individual trajectories, combining the distribution of the within-host parameters in the population (the individual model) and the infection in the households (the infection model).

#### 1. The observation probability

The observation probability was modelled as a left-censored Gaussian distribution, such that:

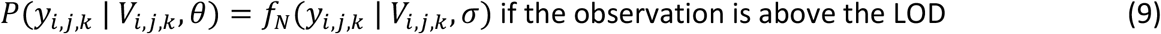

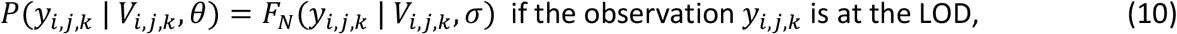

where *f*_*N*_(*y*_*i,j,k*_ | *V*_*i,j,k*_, *σ*) is the density of the normal distribution evaluated at the observation *y*_*i,j,k*_, of mean *V*_*i,j,k*_ and noise *σ*, and *F*_*N*_ is the Normal cumulative distribution function evaluated at the LOD.

#### 2. The individual viral kinetic probability

For each individual with at least one positive observation, the real viral loads *V*_*i,j,k*_ were determined by the time of infection *T*_*I,j,k*_, and by the individual parameters *T*_*P,j,k*_, *T*_*U,j,k*_, *V*_,*Pi,k*_ such that *P*(*V*_*i,j,k*_ | *θ*) = *P*(*θ*). The distribution of the individual parameters is described with

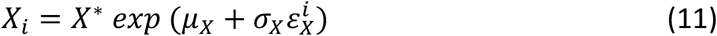

Where *X* designates the variable *T*_*P*_, *T*_*U*_, *V*_*P*_. *X*^*\**^ is the mean value of the parameter, *µ*_*x*_ is the population parameter, *σ*_*x*_ is the standard-deviation of the random effect and the 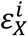 are the individual random effects. The time of infection *T*_*I,i,k*_ is inferred relatively to the reference time *T*_*min,i,k*_ = min(*T*_PCR+,*i,k*,_*T*_*sym,i,k*_), the earlier time between the first observed positive PCR test and the time of symptom onset (if declared). The time of infection is parameterized as *T*_*I,i,k*_ = *T*_*min,i,k*_ – *T*_+,*i,k*_, where *T*_+,*i,k*_ is a positive interval between infection and the reference event, that we infer jointly for each infected individual with the other parameters. For each individual and household *T*_+_ has a normal prior with location 4 days and scale 4 days, and is truncated to positive values.

#### 3. The household infection likelihood

Each infected individual contributes the probability of remaining uninfected until their infection time multiplied by the force of infection at that time. If *T*_*I,i,k*_ is the time of infection of individual *i*, their contribution to the loglikelihood is written by the sum of the two terms

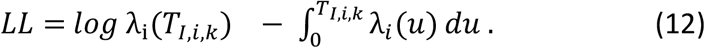

Each non-infected individual contributes the probability of remaining uninfected through the end of follow-up *T*_*f*_. The contribution to the log-likelihood of each non-infected individual is identical so that the loglikelihood is

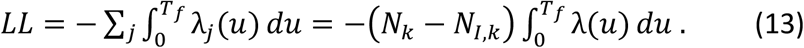

When the date of symptom onset *T*_*S,i,k*_ is available, their contribution to the likelihood is

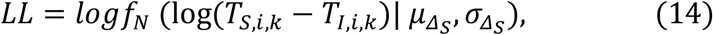

where *f*_*N*_ denotes the normal density and 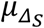 and 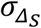 are parameters to be estimated.

**Inference in the qualitative PCR measurement design** D^PCR^

When only positivity of tests is available, the observation model is adapted from equations (9) and (10) as follows: First, the symptom onset is only available for the index case, by design, and is therefore used to infer the time of infection of the index. The observation model is a left-censored Gaussian distribution, reflecting the LOD of the PCR assay, such that

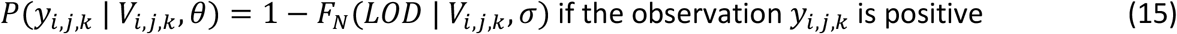

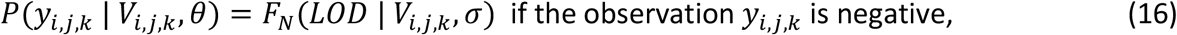

where *y*_*i,j,k*_, denotes the observed PCR result (positive or negative), *V*_*i,j,k*_ is the underlying viral load, and *F*_*N*_ is the cumulative distribution function of a normal distribution with mean *V*_*i,j,k*_ and standard deviation *σ*, evaluated at the limit of detection (LOD).

### Inference in the symptoms-only design D^TSO^

In the symptoms-only design *D*^*TSO*^, times of symptom onset were available but longitudinal virological measurements were not. Infection times were treated as latent variables and inferred from symptom-onset dates through the incubation-period distribution. Transmission was described using a parametric infectiousness profile, assumed to follow a gamma distribution. Because the full gamma distribution was weakly identifiable in this design, the rate parameter was fixed at 0.3 and only the shape parameter was estimated. The parameters estimated in this design included the transmission probability *P*_*tr*_, the household-size scaling parameter (*α*) and the shape parameter of the gamma distribution. The loglikelihood for a given household is

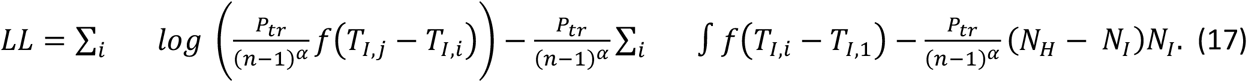

### Alternative threshold-based transmission model

As a comparison to the viral-load-dependent transmission model, we also considered a threshold-based model in which the hazard of transmission is constant whenever viral load is above the detection limit and zero otherwise. It is defined as

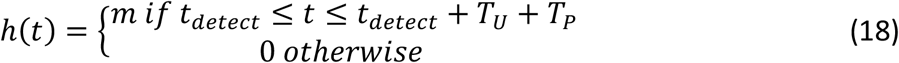

The overall transmission probability is

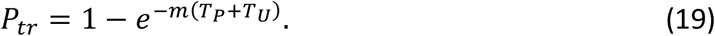

Model comparison between the viral-load-dependent and threshold-based transmission models was performed using the deviance information criterion (DIC), with models within five DIC points of the lowest value considered similarly supported by the data.

### Reconstruction of the chains of transmission

The chains of transmission were reconstructed from posterior draws of infection times, viral loads, and transmission parameters. For each infected individual, potential infectors *i* were assigned in proportion to their estimated hazards at the infection time

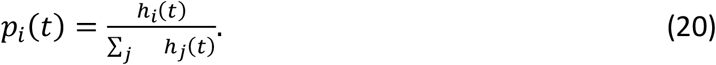

For *D*^*TSO*^ reconstruction used inferred infection times and duration of infectivity. Accuracy was assessed by the fraction of perfectly reconstructed chains and the fraction with the correct number of transmission generations. For each retained posterior parameter draw, we sampled one transmission tree by assigning to each secondary case an infector according to the relative transmission hazards at its infection time. A chain was perfectly reconstructed when every infector-infectee link matched the input tree. For each household, reconstruction accuracy was calculated as the proportion of sampled trees that exactly matched the simulated tree. These proportions were then averaged across households with at least two infections. The second measure of accuracy was the correct number of transmission generations, which quantifies that the first generation is the index case, the second generation comprises those infected by the index case, and so on. A generation number of 1 indicates that the index case infected no one else in the household. Chain reconstruction was used to estimate epidemiological parameters such as the generation interval.

## Results

### The multiscale framework reconstructs transmission chains and epidemiological parameters in an example household study

We first assessed the multi-scale modelling framework in a specific application with parameters consistent with SARS-CoV-2. Variability in transmission only arose from individual and temporal differences in viral load, producing heterogeneous infectiousness profiles and transmission events clustered near peak viral load (Figure 2B).

In the reference simulation, 100 households were simulated, generating a total of 44 infection events corresponding to a SAR of 28.0%. At least one secondary infection occurred in 37 households. The median generation interval was 5.0 days (IQR: 4.8 - 5.9 days) and the median incubation period was 4.9 days (IQR: 3.9 - 6.1 days).

With the viral load design *D*^*VL*^ the Bayesian framework reconstructed individual viral trajectories in a representative household (Figure 2C). Reconstructed chains closely matched the simulated truth (Figure 2D).

Beyond reconstructing chains of transmission, the framework also recovered distributions of epidemiological parameters. The generation interval and incubation period distributions were reconstructed with minimal bias (Fig. 2E,F). Because these quantities depend on the underlying transmission chains, their accurate recovery relies on correctly inferring who infected whom, highlighting the strength of the integrated within- and between-host approach and the fine-grained sampling. The generation interval was estimated correctly, with an average estimate of 5.4 days (95% CI: 3.6 - 7.6) across the different chains of transmission. The incubation period was estimated at 5.2 days (95% credible interval (CrI): 2.6 - 8.5). The transmission probability was slightly overestimated (Fig. 2G) with a mean posterior of 0.39 (95% CrI: 0.18 - 0.76) compared with a mean transmission probability of 0.33 across simulated individuals (95% CI: 0.12 - 0.73).

The individual parameters, infection times and viral dynamic parameters were globally well estimated in all households (Figure S1A-D). However, accuracy decreased when the ascending phase of the viral load of the index case was not observed. This occurred in 68 out of 100 index cases and resulted in systematic bias that were dependent on the input parameters, with small *T*_*P*_ being overestimated (Figure S1B) and large *T*_*U*_ and *V*_*P*_ being underestimated (Figure S1C,D). Parameter coverage was similar between index and secondary cases (Table 1).

**Table 1.** Bias and coverage obtained from the inference framework using the quantitative viral loads design *D*^*VL*^ . Coverage was estimated separately for the index case and their contacts. Values indicate the mean and 95% confidence interval across simulated datasets.

| Parameter | Description | Coverage<br>(global/index/contacts) | Bias mean (95%CI) |
| --- | --- | --- | --- |
| $T_P$ | Proliferation time | 99% / 99% / 98% | 0.059 [-0.81 - 0.86] days |
| $T_U$ | Clearing time | 97% / 96% / 98% | 0.12 [-1.4 - 0.94] days |
| $V_P$ | Peak viral load | 94% / 93% / 97% | 0.14 [-1.2 - 1.4] 0.12 log<sub>10</sub> copies/mL |
| $t_{inf}$ | Infection period | 94% / 93% / 97% | 0.07 [-1.53 - 1.69] days |

### Performance of inference across transmission probability settings and study designs

We assessed robustness of our inference framework and quantified the added value of longitudinal quantitative viral load measurements, by evaluating its performance across a range of pathogen transmissibility (*P*_*tr*_ = 15%, 30%, 45%) and the three study designs.

#### Recovery of the epidemiological parameters distributions

Across transmissibility intensities, the full framework successfully recovered key epidemiological parameters when viral load data were included. Coverage was ∼100% across all transmission probability settings, parameters (Table S2) and designs. Mean generation intervals deviated by only ∼0.2-0.3 days from the true values (Fig. 3A), incubation period by ∼0.2 days (Fig. 3B), and transmission probabilities were centered around the simulated values of 0.15, 0.30 and 0.45 (Fig. 3C).

**Figure 3.**
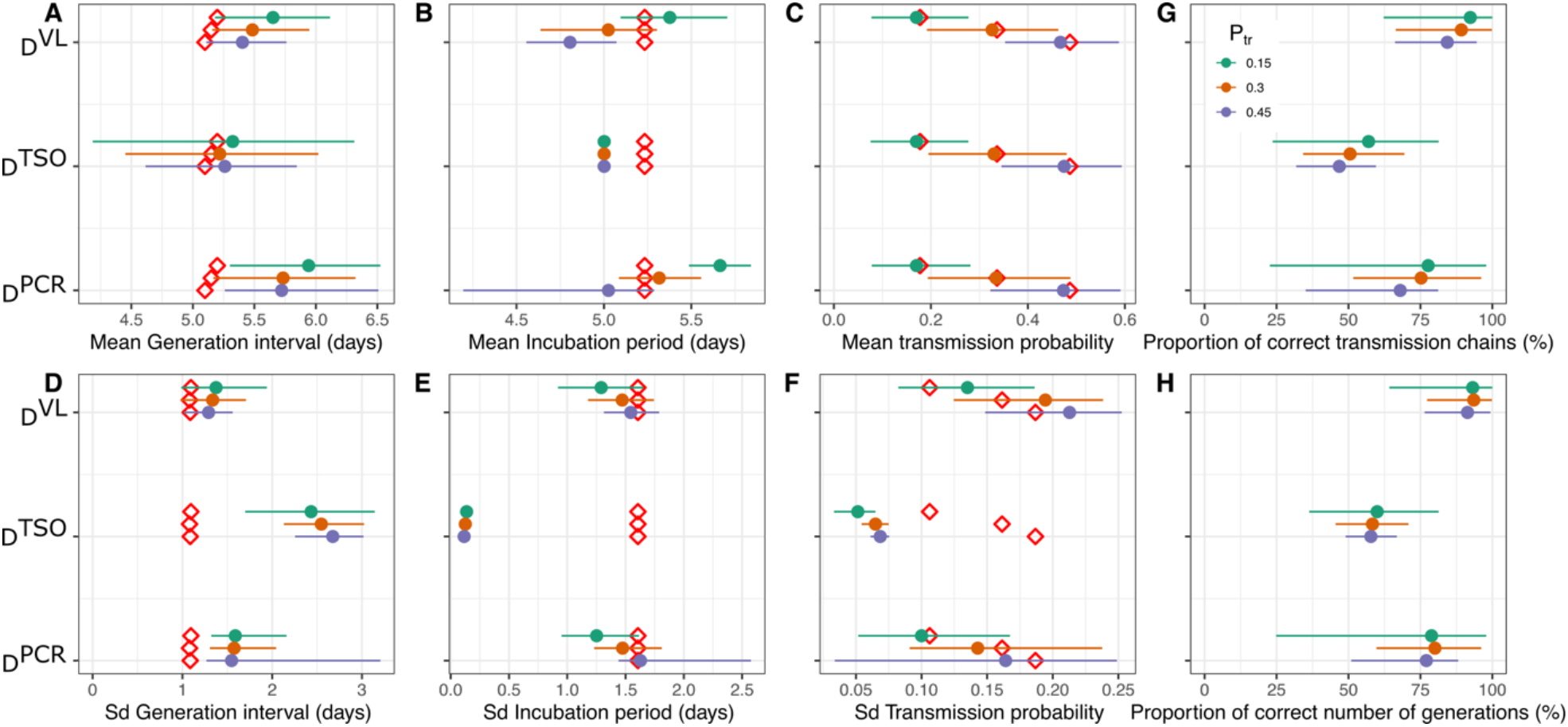
Epidemiological parameter inference for different household study designs and models. Simulations included 100 households. The mean (**A-C**) and standard deviation (sd) (**D-F**) were estimated over 100 independent repeated simulations. Results are compared across *D*^*VL*^, *D*^*TSO*^ and *D*^*PCR*^ and across input transmission probabilities *P*_*tr*_= 0.15, 0.30, and 0.45 (green, brown, blue, respectively). (**A**,**D**) Generation interval estimates, and (**B**,**E**) incubation period estimates, (**C**,**F**) transmission probability. (**G**) Proportion of correctly identified transmission chains. (**H**) Proportion of correctly estimated number of generations. Diamonds indicate true simulated values. Points and bar represent the mean and 95% CI across 100 simulation studies.

Relying only on *D*^*TSO*^ and *D*^*PCR*^ also allowed mean values to be recovered reasonably well, with bias <∼0.3 days, but uncertainty was substantially larger in *D*^*TSO*^, with credible intervals up to 2–3 times wider than in the full viral-load-based design *D*^*VL*^ (Fig. 3A-C). Crucially, considering *D*^*VL*^ also enabled the reconstruction of the variance of these parameters, not just their means. The standard deviation of the generation interval was recovered within ∼0.2-0.4 days of the true value (Fig. 3D), and that of the incubation period within ∼0.1-0.3 days (Fig. 3E). Likewise, the variance in transmission probability was faithfully reproduced (Fig. 3F), although slightly overestimated, with inferred standard deviations typically exceeding the true value by ∼0.02-0.05. In contrast, simpler data analysis designs failed to capture this variability. In design *D*^*TSO*^, all individuals shared the same infectivity profile, leading to strong underestimation or poor identification of variability in parameters. *D*^*PCR*^ performed better but still systematically underestimated standard deviations compared to the viral-load design (Fig. 3D-F). This inability to recover heterogeneity limits the interpretation of transmission dynamics, as variability in infectiousness is a key driver of epidemic spread.

#### Reconstruction of transmission chains

An essential test of the framework is whether it can accurately reconstruct the chains of transmission within households. Viral load-based design *D*^*VL*^ was accurate: between 87% and 93% of transmission chains were perfectly reconstructed depending on transmission intensity (Fig. 3G, Table S3), and the correct number of generations was recovered in 93-94% of cases (Fig. 3H, Table S3). This demonstrates that integrating within-host dynamics provides sufficient temporal resolution to infer who infected whom. Perfect reconstruction of transmission chains dropped to 46-57% with *D*^*TSO*^ and to 68-78% with *D*^*PCR*^, therefore outperformed by *D*^*VL*^. Similarly, correct identification of the number of generations decreased to ∼58-60% (*D*^*TSO*^) and ∼77-80% (*D*^*PCR*^). This limitation was particularly pronounced when symptom onset overlapped across individuals, as illustrated in the household detailed in Figure 2D, for the symptom-based (Fig. S2A) and qualitative design (Fig. S2B).

### Quantitative viral load data enables recovery of the association between viral load and transmissibility

We first assessed whether the framework accurately recovered the transmission parameter *m* across transmission probability settings. For each simulated dataset, we summarized the posterior distribution of *m* by its posterior median. Across all transmission intensities, estimates obtained with the viral-load-based design were centered around the true input values, indicating that the model successfully captured the overall level of transmission (Fig. 4A). Design *D*^*PCR*^ also recovered the transmission parameter reasonably well, although with increased variability. In contrast, *D*^*TSO*^ provided substantially less information on transmission heterogeneity because all infected individuals shared the same infectiousness profile.

**Figure 4.**
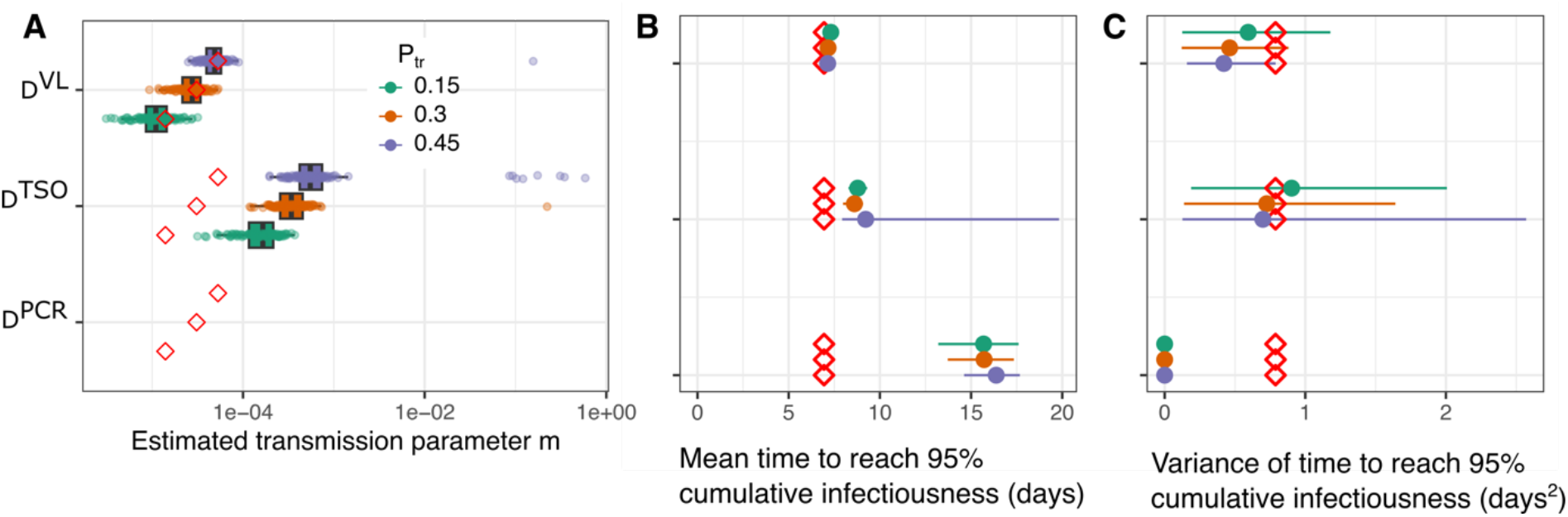
Recovery of transmission parameters and infectiousness profiles across household study designs. (**A**) Recovery of the transmission parameter *m*. Boxes show the distribution of posterior medians of *m* across 100 simulated datasets for each input transmission probability. Red diamonds indicate true input values. (**B**) Mean time required to reach 95% of cumulative infectiousness across the three study designs. (**C**) Variance of the time required to reach 95% of cumulative infectiousness. Colors indicate the input transmission probability (*P*_*tr*_= 0.15, 0.30, 0.45). Points represent the mean estimate across simulations and horizontal bars the 95% confidence intervals. Red diamonds indicate the corresponding values obtained from the input transmission model.

We then compared the inferred temporal profiles of infectiousness across the three study designs by evaluating the distribution of the time required to reach 95% of cumulative infectiousness (Fig. 4B-C). This quantity is the first time *T*_95_ such that 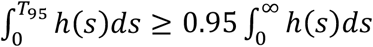 and summarizes how rapidly transmission potential accumulates following infection. The design based on viral load accurately recovered both the mean and the variance of this distribution across transmission scenarios, closely matching the simulated values. The qualitative PCR design systematically overestimated the time required to reach 95% infectiousness and showed inflated uncertainty, particularly at higher transmission probabilities. By construction, the symptom-based design inferred a single infectiousness profile shared across individuals, resulting in negligible estimated variance.

## Discussion

Using simulations, we demonstrate that integrating quantitative viral load data into household transmission studies improves inference of epidemiological parameters, transmission chains, and individual heterogeneity in infectiousness. Although designs based on time of symptom onset or qualitative PCR recovered mean parameters reasonably well, they provided limited information on the timing and variability of individual infectiousness. As a result, they failed to reliably recover parameter distributions and inter-individual heterogeneity. Longitudinal viral load measurements partly overcome these limitations by providing individual-level temporal information on viral trajectories, enabling recovery of parameter distributions rather than averages alone.

The added value of viral load data persisted even under favorable assumptions for symptom-based designs. In a supplementary analysis assuming perfect knowledge of symptom onset for all individuals (Fig. S5), inference results remained very similar to those obtained in the main analysis (Fig. 3) where symptoms were not included. This indicates that the improvement primarily arises from the quantitative information contained in viral load trajectories rather than from improved knowledge of symptom timing alone. This is consistent with studies showing that viral load is associated with transmission risk [9,10] and that infectiousness varies over the course of infection, often around periods of high viral shedding [16,17].

In our simulations, the framework reconstructed individual infectiousness profiles and variability in transmission-related parameters when transmission was driven by viral kinetics (Figs. 3D–F and 4B). Because heterogeneity in infectiousness is a major determinant of epidemic spread and even superspreading potential [18,19], failing to account for it may lead to mischaracterization of transmission dynamics. Although viral load is used here as a primary driver of infectiousness, our framework can accommodate other sources of heterogeneity. In an additional analysis (Fig. S4), we introduced structured heterogeneity by assigning individuals to categories with different transmission parameters. The model successfully recovered these category-specific effects, demonstrating its ability to capture covariate-driven differences in infectiousness beyond viral kinetics alone.

Several limitations should be acknowledged. First, the framework was evaluated exclusively using simulated household outbreaks. This allowed reconstruction accuracy to be assessed against known infection times and transmission chains. However, the simulated outbreaks were generated under the same viral-kinetic and transmission assumptions used for inference. The results therefore demonstrate performance under these assumptions, rather than validating the assumed relationship between viral load and infectiousness. Evaluation using observed household data and assessment of robustness to model misspecification are needed to establish practical performance. Second, simulations relied on a simplified household model assuming homogeneous contacts and no community-acquired infections. Relaxing these assumptions by incorporating community transmission, heterogeneous susceptibility, or alternative transmission mechanisms such as cumulative exposure would improve realism and should be explored in future work. Third, quantitative PCR measures viral RNA rather than infectious virus, and the relationship between RNA viral load, culture positivity, and transmissibility may vary across individuals and over the course of infection [20,21]. For example, Vargas-Maldonado et al. [22] reported associations between infectious virus expulsion, respiratory viral load, and symptoms, alongside substantial individual heterogeneity. These findings highlight the limitations of representing infectiousness solely through a fixed function of measured viral load. The square-root relationship used here is a modelling assumption. Future work should assess alternative relationships and allow for additional determinants of infectiousness.

Finally, the present implementation was designed to mimic SARS-CoV-2 infection, but the hierarchical framework linking individual infection trajectories to household transmission could be adapted to other respiratory pathogens. Such applications would require pathogen-specific viral kinetics, observation models, and relationships between measured viral load and infectiousness. For RSV, a longitudinal household study found that symptomatic index cases were more likely to be associated with household spread than asymptomatic index cases. Higher peak nasopharyngeal viral load and longer shedding duration were also associated with increased odds of spread [23]. Studies of influenza and SARS-CoV-2 have linked respiratory viral load and symptoms to infectious virus expulsion [22] and aerosol RNA shedding [24], respectively. However, substantial individual heterogeneity in influenza and differences across SARS-CoV-2 variants and specimen types highlight the limitations of using sampled viral load alone to characterize infectiousness. Together, these findings support considering viral kinetics as one component of transmission models, while emphasizing that the relationship between sampled viral load and infectiousness requires empirical assessment for each pathogen.

Although this study focused on households, the framework could be extended to other structured environments such as schools, workplaces, or long-term care facilities. In these settings, integrating biological, behavioral, and environmental determinants of transmission will likely be essential. Overall, our results support the integration of quantitative viral load measurements into future transmission studies and provide quantitative guidance for the design of studies aiming to better characterize infectious disease transmission dynamics.

## Data Availability

The code generated for the current study is available in the GitHub repository: https://github.com/nathoze/households and the simulated data is in the Zenodo repository 10.5281/zenodo.20702104.

https://zenodo.org/records/20702104

