## Supplementary Information for "A multiscale mathematical model integrating viral load improves reconstruction of transmission chains and epidemiological parameters in household studies"

^*^ Dual lead authorship

***Implementation***

We estimated simultaneously the individual parameters of the viral kinetics $T_{P}, T_{U}, V_{P}$the times of infection $T_{I}$and the population transmission parameter $m$. We used rstan to estimate the parameters using a Markov Chain Monte Carlo (MCMC) algorithm [1]. The No-U-Turn sampler variant of Hamiltonian Monte Carlo was used to update the parameters and the runs are 3000 iterations, the first 1500 being warmup. The priors for the different parameters are for the population parameters $N\left( 0,0.25 \right)$. For the random effects they are $N\left( 0,0.25 \right) | \sigma_{T_{P}}>0$.

We allow the household-size scaling parameter $\alpha$to take negative values (corresponding to effective per-contact transmission increasing with household size) and gave it a normal prior $N\left( 0.5,1 \right).$ The residual noise $\sigma\sim N\left( 0.5,0.1 \right)| \sigma>0$. The transmission parameter is non informative in the logarithmic scale such that $log\left( m \right) \sim N\left( -5,3 \right)$. In the constant model we chose the same prior except for the transmission parameter which is $log\left( m \right) \sim N\left( -3,3 \right)$.

For each individual *i,* $T_{+,i} \sim N\left( 4,4 \right) | T_{+,i}>0$*.*

For the design $D^{TSO}$, non-informative priors are chosen for the shape parameter of the infectious period distribution$\gamma\sim N\left( 10,10 \right)| \gamma>0$, for the infection probability with $log \left( P_{tr} \right) \sim N\left( -3,3 \right)$ and for the population parameter of the incubation period $\mu_{\Delta_{S}}\sim N\left( 3,3 \right)| \gamma>0.$

***Accounting for covariates of infectiousness***

To evaluate the ability of the framework to recover covariate-driven heterogeneity, we simulated populations with two categories of infectiousness defined by different transmission parameters. These categories were defined by two different values of the transmission strength parameter *m*, while viral dynamics parameters were otherwise unchanged. We generated 100 independent datasets, each consisting of 100 households, with individuals equally distributed between categories corresponding to average individual transmission probabilities of 30% and 50%. We fitted the model to each dataset assuming knowledge of the categorical covariate.

Across simulations, the framework successfully recovered both transmission strength parameters. The distribution of posterior mean estimates of *m* for each category was centered close to the corresponding input values, with limited overlap between categories (Figure S4). Moreover, in all simulations, credible intervals contained the true values, demonstrating reliable identification of covariate effects.

***Identification of the viral load-infectiousness relationship***

We next assessed whether the study design enables identification of the functional relationship between viral load and transmission. To do so, we evaluated whether the inference framework could recover the correct underlying mechanism by fitting an alternative transmission model to the same simulated datasets. The competing model differed in how transmissibility depended on viral load (threshold-based transmission vs. peak-dependent transmission in the baseline model). Model comparison was performed using the deviance information criterion (DIC). The true transmission model was correctly identified in 89% of simulations, demonstrating that the study design provides sufficient information to discriminate between competing hypotheses on the relationship between viral load and infectiousness. Increasing the number of households further improved model identification (approaching 95-100% in larger samples, Fig. S3), particularly at higher transmission probabilities. Identification improved at higher transmission probabilities, likely because increased transmission generates longer and more informative chains, enhancing the ability to resolve temporal patterns in infectiousness.

**Supplementary figures**

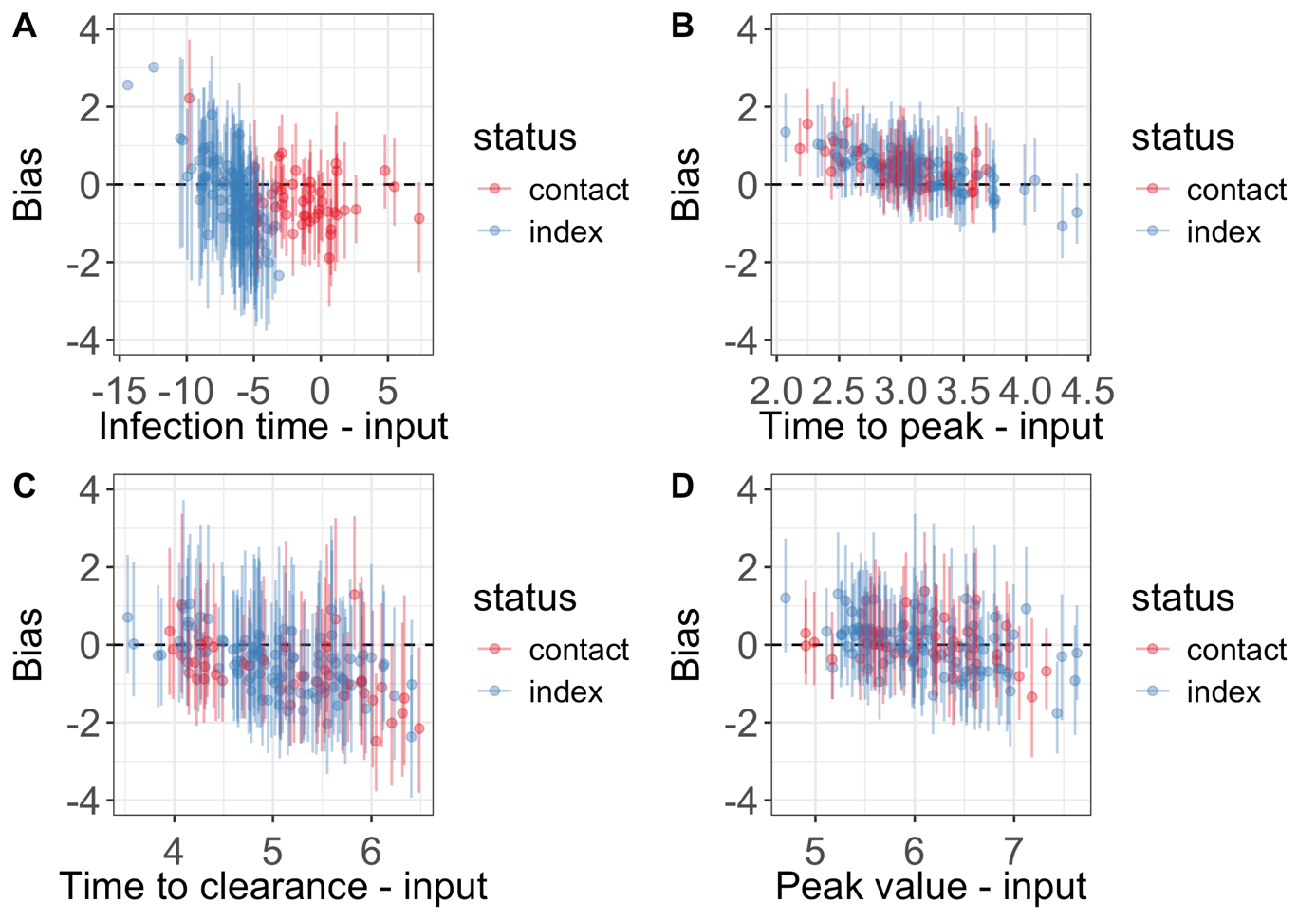

**Figure S1: Estimation of individual parameters in a simulated dataset.** (A) Bias in inferred infection times, showing posterior means and 95% credible intervals. Time is expressed relative to household enrollment (time 0). (B-D) Bias in posterior estimates of the time to peak viral load $T_{P}$ (B), time to clearance $T_{U}$ (C), peak viral load $V_{P}$ (D). Blue indicates the index case, red one of the contacts.

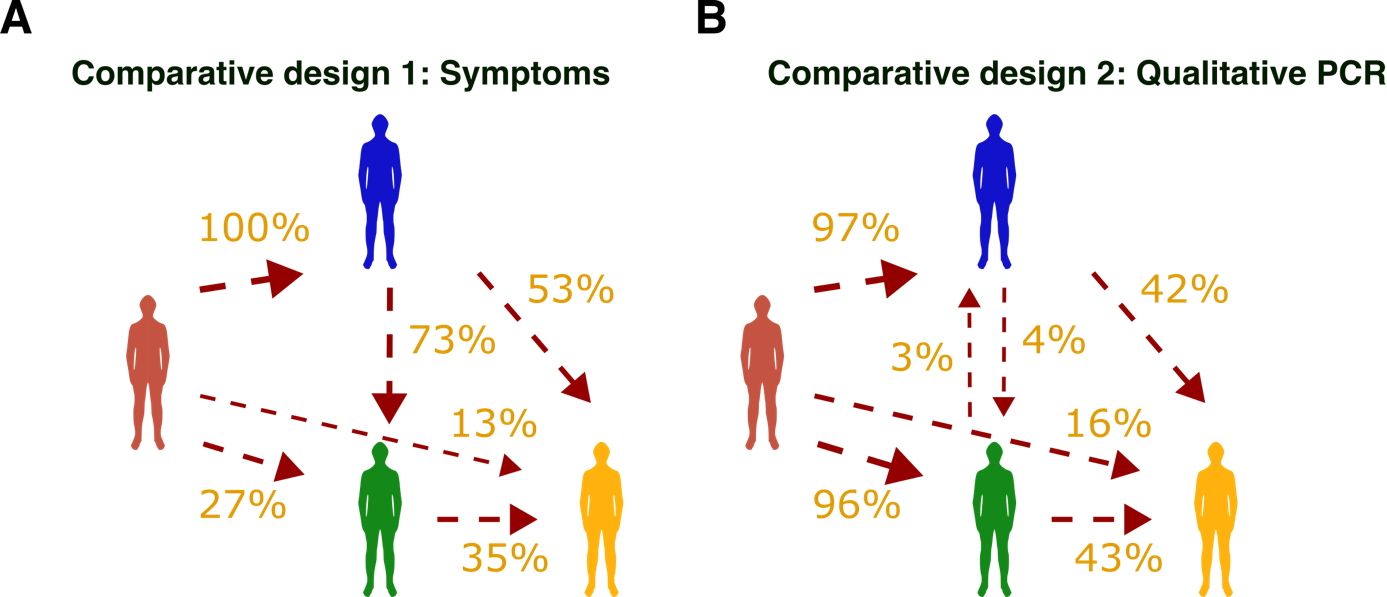

**Figure S2. Transmission chains reconstruction using alternative study designs.** Reconstruction of transmission chains in the representative household shown in Figure 2D using (A) $D^{TSO}$ and (B) $D^{PCR}$.

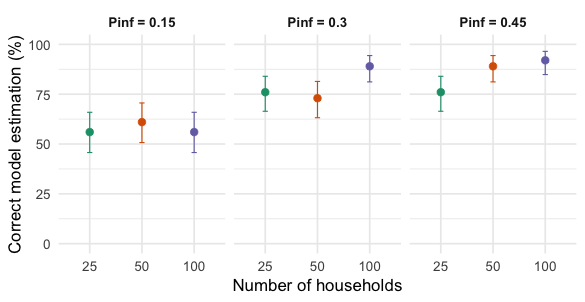

**Figure S3.** **Identification of the correct model of virus transmission**. Comparison was assessed using the Deviance Information Criterion (DIC) over 100 simulated datasets. Vertical bars denote 95% binomial confidence intervals.

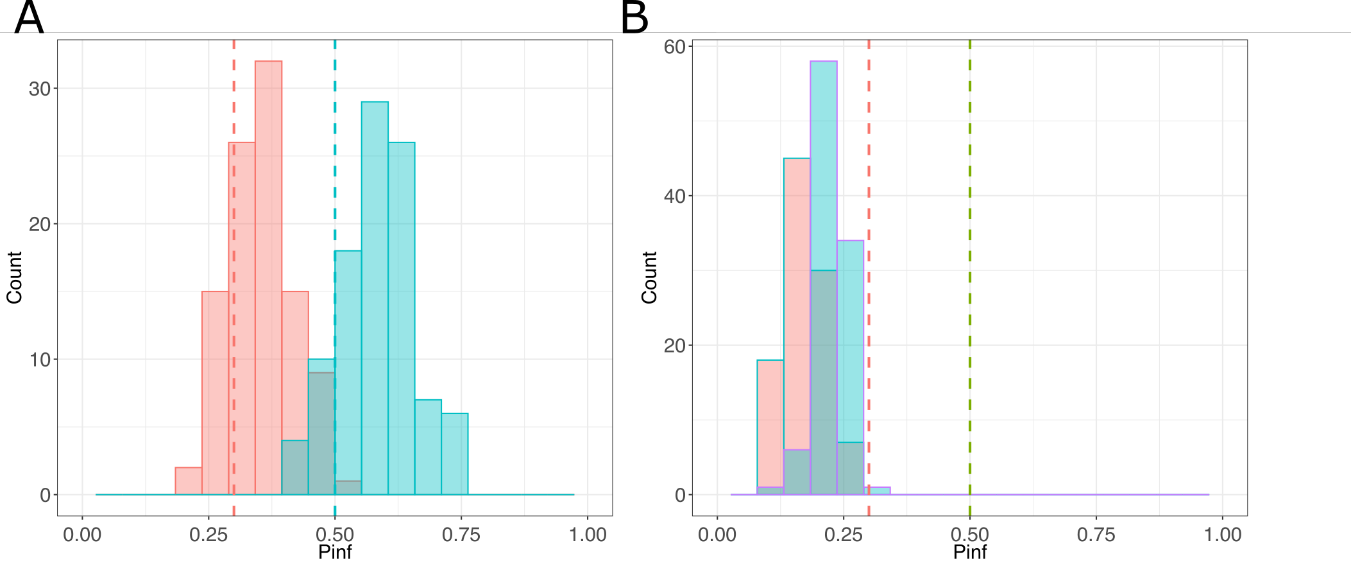

**Figure S4**. Recovery of infectiousness covariate effects. Distribution of posterior mean estimates of the transmission strength parameter *m* across 100 independent simulation replicates (each replicate includes 100 households), under a model with two infectiousness categories. Individuals were assigned to category 1 or 2, corresponding to two distinct values m_1_ and m_2_. Histograms show the distribution (over the 100 replicates) of the estimated *m* for each category. Vertical dashed lines indicate the true input values used to generate the data.

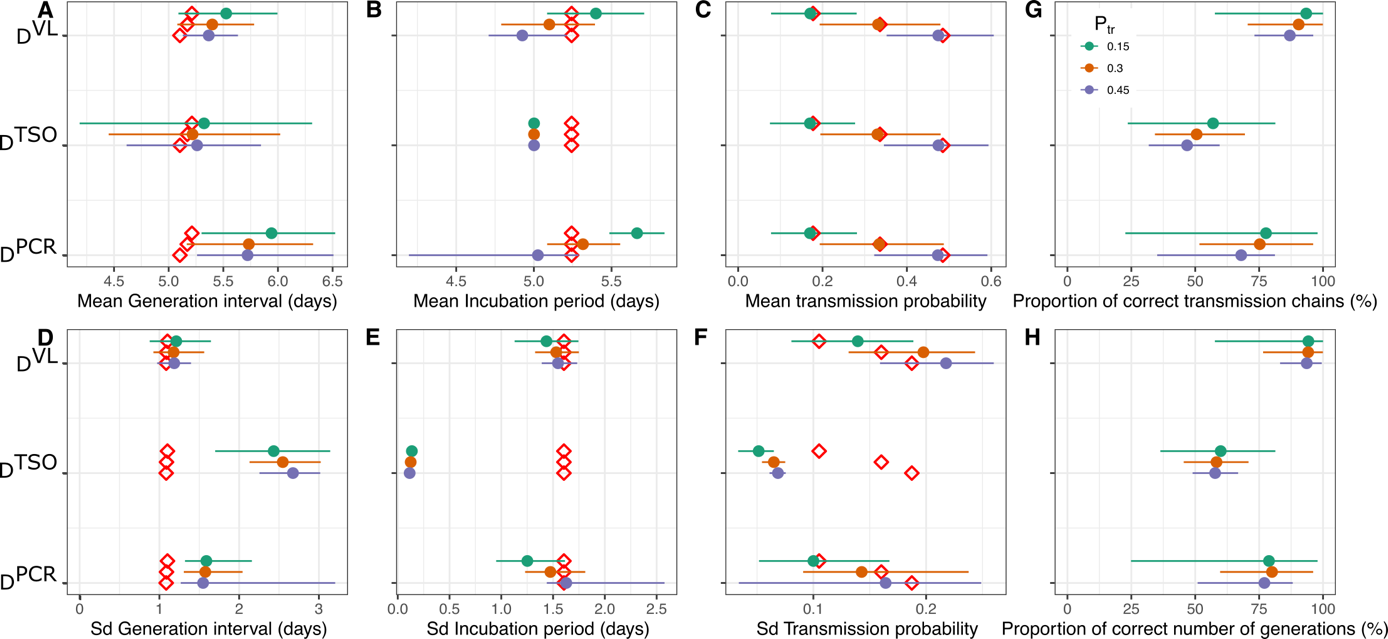

**Figure S5**. Recovery of epidemiological parameters under the assumption that symptom onset dates are available for all infected household members.

**Table S1**. Parameter values used to generate individual viral-dynamics profiles and household transmission in the simulations.

| **Parameter** | **Description** | **Value** | **Sd of the random effects** |
| --- | --- | --- | --- |
| $T_{P}$ | Proliferation time | 3 days | 0.15 |
| $T_{U}$ | Clearing time | 5 days | 0.15 |
| $V_{P}$ | Peak viral load | 6 log_10_ copies/mL | 0.1 |
| $\Delta_{S}$ | Incubation period | 5 days | 0.3 |
| $\alpha$ | Household-size scaling parameter | 1 | - |
| $\sigma$ | Observation noise | 0.8 | - |
| $P_{tr}$ | Transmission probability | 0.15, 0.30, 0.45 | - |

**Table S2**. Coverage of inferred parameters in the simulation across transmission-probability settings in the three data collection designs. Values indicate the percentage of 100 simulated datasets in which the input value was contained in the 95% credible interval. Inferred parameters are the generation interval (GI), the incubation period, the transmission probability ($P_{tr}$). The three designs are indicated in parenthesis.

| $P_{tr}$ | **GI (**$D^{VL}$**)** | **GI (**$D^{TSO}$**)** | **GI (**$D^{PCR}$**)** | **Incubation (**$D^{VL}$**)** | **Incubation (**$D^{TSO}$**)** | **Incubation (**$D^{PCR}$**)** | $P_{tr}$ **(**$D^{VL}$**)** | $P_{tr}$ **(**$D^{TSO}$**)** | $P_{tr}$ **(**$D^{PCR}$**)** |
| --- | --- | --- | --- | --- | --- | --- | --- | --- | --- |
| 0.15 | 100% | 100% | 100% | 100% | 100% | 100% | 100% | 92% | 99% |
| 0.3 | 100% | 100% | 100% | 100% | 100% | 100% | 100% | 91% | 98% |
| 0.45 | 100% | 100% | 100% | 100% | 100% | 100% | 100% | 94% | 95% |

**Table S3**. Accuracy of transmission chain reconstruction in the different data collection designs and transmission strength. Values correspond to Figure 3G-H and indicate the fraction of perfectly reconstructed chains and the fraction of households for which the correct number of transmission generations was recovered. $P_{tr}$: Input transmission probability in the simulations. The three designs are indicated in parenthesis.

| $P_{tr}$ | **Full chains (**$D^{VL}$**)** | **Full chains (**$D^{TSO}$**)** | **Full chains (**$D^{PCR}$**)** | **Number of generations**  **(**$D^{VL}$**)** | **Number of generations (**$D^{TSO}$**)** | **Number of generations (**$D^{PCR}$**)** |
| --- | --- | --- | --- | --- | --- | --- |
| 0.15 | 93.4% | 57.0% | 77.7% | 94.3% | 60.0% | 78.9% |
| 0.3 | 90.5% | 50.6% | 75.2% | 94.2% | 58.3% | 80.0% |
| 0.45 | 87.0% | 46.8% | 68.0% | 93.6% | 57.8% | 77.1% |
